# Capillary angiopathy and aquaporin-4 after Aβ immunisation in Alzheimer’s disease – potential relevance to Amyloid-Related Imaging Abnormalities

**DOI:** 10.1101/2022.12.08.22283144

**Authors:** Charlotte H. Harrison, Kenji Sakai, David A. Johnston, Clive Holmes, Delphine Boche, James AR Nicoll

## Abstract

**Aims:** Amyloid-related imaging abnormalities (ARIA) have hampered clinical trials and therapeutic use of amyloid-β (Aβ) immunotherapy for Alzheimer’s disease (AD), with the cause of the white matter oedema (ARIA-E) unknown. Aquaporin 4 (AQP4), present in astrocyte endfeet, controls water flow across the blood-brain barrier. Experimental studies suggest that as Aβ plaques are cleared following immunotherapy, capillary angiopathy (capCAA) increases, displacing astrocyte endfeet allowing influx of extracellular water (oedema). We sought neuropathological evidence for this mechanism in immunised AD patients.

**Methods:** Brains of 16 Alzheimer’s patients immunised against Aβ42 (iAD, AN1792, Elan Pharmaceuticals) and 28 unimmunized Alzheimer’s (cAD) cases were immunolabelled and quantified for Aβ42 and AQP4.

**Results:** CapCAA was 3.5 times higher in iAD (p=0.009). No difference between the groups was identified in the proportion of capillaries wrapped by AQP4 or AQP4 protein load. However, capCAA in iAD negatively correlated with AQP4 load (r = -0.498, p<0.001), suggesting disturbance of AQP4 in presence of capCAA.

**Conclusions:** After Aβ immunotherapy, capCAA was increased, likely reflecting the drainage of soluble Aβ towards the vasculature and providing a potential mechanism to disrupt AQP4-containing astrocyte endfeet, resulting in ARIA-E. We did not identify alterations in AQP4, potentially because of limitations in the timing of the post-mortem analysis. Given the recent licencing of Aβ immunotherapy, the field must prioritise obtaining neuropathological correlates of ARIA to explore its mechanisms further.

## Introduction

The amyloid cascade hypothesis [1] stipulates that amyloid-β (Aβ) aggregation is the initiating event in AD. The hypothesis has been tested therapeutically in a range of active and passive immunotherapy trials targeted against Aβ [2]. The first clinical trial, using an active anti-Aβ42 vaccine (Elan Pharmaceuticals AN1792) [3] failed to demonstrate slowing of cognitive decline [4], while *post-mortem* findings showed clearance of plaques [4-7] and exacerbation of cerebral amyloid angiopathy (CAA) [8, 9]. This was attributed to plaques being disaggregated by anti-Aβ antibodies [10] and phagocytic microglia [11], permitting Aβ to enter the perivascular drainage pathway [12-14] and be removed from the brain [8, 15].

Subsequent Aβ immunotherapy trials incorporated systematic brain imaging into their study design [16], allowing the identification of amyloid-related imaging abnormalities (ARIA) as radiological observations interpreted as representing microhaemorrhages (ARIA-H) and oedema (ARIA-E) [16-18]. ARIA has become a persistent obstacle in Aβ immunotherapy trials [2, 19], its presence limiting dosing regimens and curtailing clinical trials, hampering completion of studies to assesses efficacy. ARIA is associated with possession of the apolipoprotein ε4 allele (APOE4), higher antibody doses and localised reduction of amyloid assessed in vivo by PiB PET scans [17, 20]. ARIA-H has been proposed to be due to increased severity of CAA following immunotherapy causing microbleeds [8, 15]. ARIA-E is defined as radiological hyperintensity in the grey or white matter or leptomeninges of the brain, which represent oedema [16], in particular seems to be restricted to the weeks after onset of treatment and is reversible. From a pathophysiological point of view, vascular permeability [8, 16], inflammation [15, 21, 22] and APOE isoform [16] have been proposed to play a role in causing ARIA-E; however the exact mechanism remains unknown.

Of the two main pathogenic isoforms of Aβ, Aβ42 is the main constituent of parenchymal plaques and capillary cerebral amyloid angiopathy (capCAA) located within the capillary basement membrane [23, 24]; whereas Aβ40 predominates in the walls of arteries and arterioles, forming CAA [24]. CAA is classified into two distinct histopathological phenotypes, with the presence of capCAA and the possession of an APOE4 allele distinguishing type 1 CAA from type 2 [25, 26]. Unlike arterial and arteriolar CAA, capCAA may be surrounded by tau deposits and reactive microglia [27] and results in functional blood flow disturbances, which may exacerbate Aβ-mediated neurotoxicity [28]. Therefore, although capCAA is typically an infrequent feature of AD [29, 30], it may be biologically important in its pathogenesis. In PDAPP mice, capCAA was increased after passive Aβ immunisation [31], and although capCAA has been noted to occur in human AD following Aβ immunotherapy, it has not been systematically studied or quantified [6, 9].

Aquaporin-4 (AQP4), the principal water transporter in the CNS [32, 33], offers a potential link between the vascular Aβ changes following immunisation and ARIA-E. AQP4 is expressed by astrocytes [34] at interfaces between brain parenchyma and fluid, most notably in perivascular astrocyte endfeet [35-37], and is integral to maintain astrocyte function, including fluid and ion homeostasis [33, 38, 39]. Experimental [31, 40-42] and human studies [40] have demonstrated a relationship between Aβ deposits and AQP4 distribution. In areas of vascular Aβ, AQP4 is redistributed away from perivascular endfeet, with a concurrent clustering of AQP4 around parenchymal plaques [43-46]. In the PDAPP mouse model immunised against Aβ, AQP4 was reduced in areas of newly formed capCAA. In addition, there was structural disorganisation of astrocyte endfeet [31], displacing endfeet away from their normally tight contact with endothelial cells, potentially allowing efflux of water from the blood into the brain parenchyma, causing extracellular water accumulation. Images suggested a similar mechanism may occur in human AD [31]. However, no such study has been performed to date in the brains of AD patients immunised against Aβ and this might provide important insights into the pathophysiology of ARIA-E. We therefore hypothesise that an increase in capCAA may occur following Aβ immunisation and cause displacement of astrocyte AQP4-containing endfeet away from the vessel wall, or relocation of AQP4 within astrocytes away from the endfeet, leading to impaired water transfer across the capillary and resulting in interstitial oedema (ARIA-E).

In this study, we took the opportunity offered by our unique neuropathological cohort of immunised AD cases to explore certain components of the aforementioned hypothesis. Specifically, by assessing whether Aβ removal was associated with (i) increased capCAA (ii) decreased AQP4 expression and (iii) displacement of AQP4-containing astrocyte endfeet away from capillaries.

## Materials and Methods

### Characteristics of the cases

Clinical and neuropathological follow-up of Alzheimer’s patients (age range 63-89 years) enrolled in the Elan Pharmaceuticals phase 1 trial of AN1792 was previously reported [3, 4, 7]. Patients (or carers) were invited to consent to *post-mortem* neuropathological examination, and subsequently, tissue was available from 22 immunised patients with 16 having a neuropathological diagnosis of AD (termed iAD) (Table 1). Six had another cause of dementia and were excluded from further analysis. One patient (case no. 1) required imaging in life, which demonstrated features of meningoencephalitis and neuroradiological features consistent with the later defined ARIA-E [5, 16]. The case IDs were not known to anyone outside the research group.

**Table 1:** Demographic information on the immunised and non-immunised AD groups.

| Case no.<br>(iAD) | Sex | Dementia<br>duration<br>(years) | APOE<br>status | Mean<br>antibody<br>response<br>(ELISA units) | Survival time from<br>first AN1792<br>immunisation<br>(months) |
| --- | --- | --- | --- | --- | --- |
| 1** | F | 6 | 3.4 | 1:119 | 20 |
| 2 | M | 11 | 3.3 | <1:100 | 4 |
| 3 | M | 6 | 3.3 | <1:100 | 41 |
| 4 | F | 10 | 3.3 | 1:4072 | 44 |
| 6 | M | 7 | 3.4 | 1:1707 | 57 |
| 7 | M | 6 | 3.4 | 1:4374 | 60 |
| 8 | M | 10 | 3.4 | 1:6470 | 64 |
| 9 | M | 11 | 4.4 | 1:491 | 63 |
| 10 | F | 11 | 3.3 | 1:137 | 86 |
| 11 | M | 12 | 3.4 | 1:142 | 94 |
| 16 | F | 15 | 3.4 | 1:142 | 111 |
| 17 | F | 13 | 4.4 | <1:100 | 141 |
| 19 | F | 19 | Unknown | 1:221 | 162 |
| 20 | M | 17 | Unknown | 1:430 | 166 |
| 21 | F | 18 | 3.4 | 1:3045 | 173 |
| 22 | M | 18 | 4.4 | 1:1313 | 184 |
| cAD<br>(n=28) | 15 F: 13 M | 3-17 | 21 ε4+<br>6 ε4-<br>1 unknown | n/a | n/a |

\*\* Case with ARIA-E; iAD: immunised Alzheimer's case; cAD: non-immunised Alzheimer's case; F: Female; M: Male; n/a: not applicable

As there were inadequate numbers of *post-mortem* placebo treated samples from the original trial, 28 unimmunised AD cases used as controls (cAD; age range 63-88 years) were sourced from the South West Dementia Brain Bank. Cases were matched as closely as possible for age at death, APOE genotype and sex.

### Immunohistochemistry

Four μm-thick sections of formalin fixed paraffin-embedded tissue were immunostained for Aβ42 (clone 21F12, 1:4000, Prothena Biosciences). Sections from three neocortical areas – the medial frontal lobe, middle temporal lobe and inferior parietal lobule – were immunostained for AQP4 (H-80, Santa Cruz, Dallas, USA, 1:1000) and Aβ42 (Clone 21F12, Elan Pharmaceuticals). Immunolabelling was performed using the appropriate antigen retrieval method, and the signal was amplified via the avidin-biotin-peroxidase complex method (Vectastain Elite) for a detection with 3,3’-diaminobenzidine as the chromogen (Vector Laboratories, Peterborough, UK). All experiments included a negative control slide incubated in buffer with no primary antibody, and a positive control slide.

### Quantification

Slides stained for Aβ42 and AQP4 were scanned using Olympus VS110 slide scanner at magnification x40 (CapCAA, AQP4 endfeet) or x20 (Aβ42 and AQP4 loads). Olympus VS-Desktop software (v2.9) was used to extract regions of interest (ROIs) with the quantification performed in each neocortical region and blind to immunisation status.

#### CapCAA

Thirty-five ROIs of 1.58mm^2^ each were placed contiguously in the area of cerebral cortex sampled in adjacent slides for Aβ42 and AQP4. Identification of a capillary required an identifiable endothelial nucleus and/or cuboidal erythrocytes in the lumen, and a diameter less than 10μm. The number of capCAA vessels was counted in each ROI.

#### AQP4 capillary endfeet

The number of capillaries surrounded by AQP4-positive endfeet (identified by the presence of AQP4 staining surrounding the capillary outer wall) were counted in 15 ROIs placed contiguously.

#### Aβ42 and AQP4 loads

Images of thirty ROIs were captured in a zigzag manner to sample the whole thickness of the neocortex. The percentage area of Aβ42 or AQP4 staining was measured using Image J 1.45v software and expressed as protein load (%) as previously reported [9].

### Statistical analysis

Statistical analysis was performed in SPSS (ISM v25) and graphs were generated using GraphPad (PRISM v8). Due to the highly patchy nature of plaque removal observed in the immunised cohort, data from the three neocortical areas were analysed and plotted individually for each parameter used. Normality of the data was assessed using one-sample Kolmogorov-Smirnoff tests and through examination Q-Q plots. Due to the non-parametric distribution of the data, Mann-Whitney U-test was performed for group comparison. For correlations Pearson’s or Spearman’s rank test, based on the data distribution, was done within each group to determine the relationships between capCAA, AQP4 and Aβ42, and in relation with the duration of dementia, age at death, and for the iAD group with the immune response (antibody peak and mean) and the survival time after immunotherapy. Statistical tests were conducted at the 5% for intergroup comparisons and 1% significance level for correlations.

## Results

### CapCAA

CapCAA was defined as Aβ42 located within or in close association with the capillary wall. Examples of an unaffected capillary and capCAA in longitudinal and transverse section are illustrated in Figure 1A-C. CapCAA can be an early consequence of Aβ immunotherapy as it was prominent in association with disrupted plaques in one patient as little as 4 months after the first immunisation dose (Figure 1D). In addition, capCAA in areas of cerebral cortex devoid of plaques was notable in several long-term survivors, up to 14 years after immunisation, indicating it to be a longstanding and/or late recurrent feature (Figure 1E-G).

**Figure 1.**
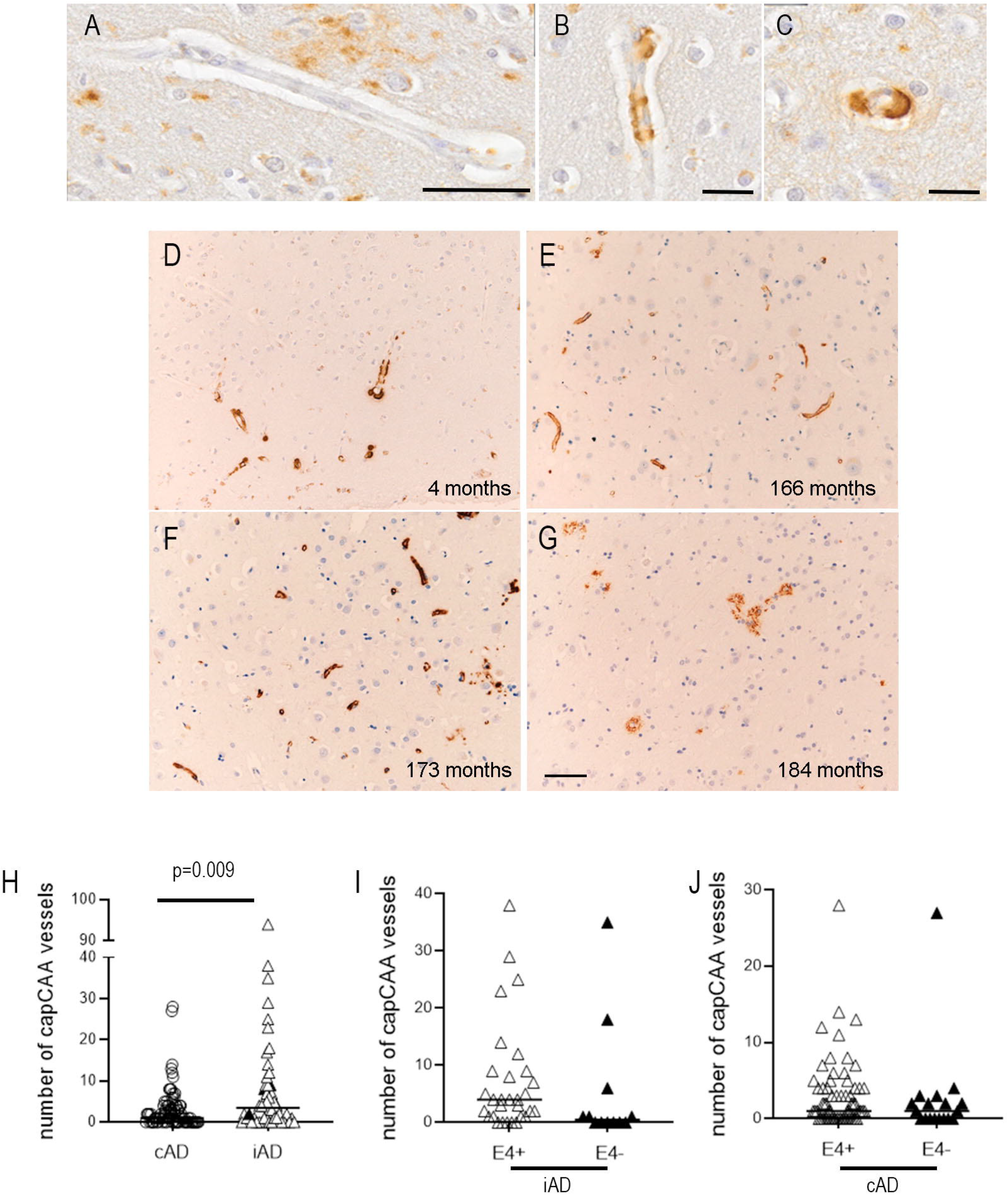
Illustrations of cerebral cortical capillaries unaffected (A) and affected (B and C) by accumulation of Aβ within and in close association with their walls (i.e. capillary CAA). Prominent capillary CAA was detected as early as 4 months after first Aβ immunisation dose in one case (case no. 2) during what appeared to be a dynamic phase of plaque removal (D). In addition, in some long term survivors, marked capillary CAA in the absence of plaques was present in some cortical regions up to 14 years after immunisation indicating it could also be a longstanding and/or late recurrent feature (E, case no. 20; F, case no. 21; G, case no. 22, Aβ42 immunohistochemistry). Quantification showed significantly more capCAA in the cerebral cortex of immunised AD cases compared with untreated AD cases (H). The case known to have had ARIA (case no. 1) is indicated in red. Amongst the immunised AD cases, there was a trend for more capCAA in *APOE* ε4 carriers than non-ε4 carriers (p=0.056) (I). No such trend was evident in *APOE* ε4 carriers in untreated AD cases (J). Scale bar (A-C) = 50μm; (D-G) =.100μm.

Quantification of capCAA showed significantly more capCAA in the iAD *vs* cAD cases (median iAD=3.5 *vs* cAD=1, p=0.009; Figure 1H). In order to determine the influence of *APOE* ε4 on capCAA, subgroup analysis was performed according to *APOE* ε4 allele status (ε4 carriers *vs* non-carriers). In iAD, there was eight times more capCAA in ε4 carriers compared to non-ε4 carriers, but with borderline significance (median ε4+=4 *vs* ε4-=0.5, p=0.056; Figure 1I). In cAD, there was no difference in capCAA based on ε4 status (p=0.238; Figure 1J).

### AQP4

AQP4 expression was detected in the cell bodies and processes of astrocytes (Figure 2A). A patchy staining pattern of AQP4 was noted in the cerebral cortex, likely reflecting astrocytes associated with Aβ plaques, as previously described. There was dense AQP4 immunolabelling in astrocytes located in the subpial cortex and in the periventricular subependymal region. Variable AQP4 expression was observed around large CAA-affected blood vessels with more intense staining attributed to AQP4-positive astrocyte endfeet around some, but not all, capillaries (Figure 2B, 2C).

**Figure 2.**
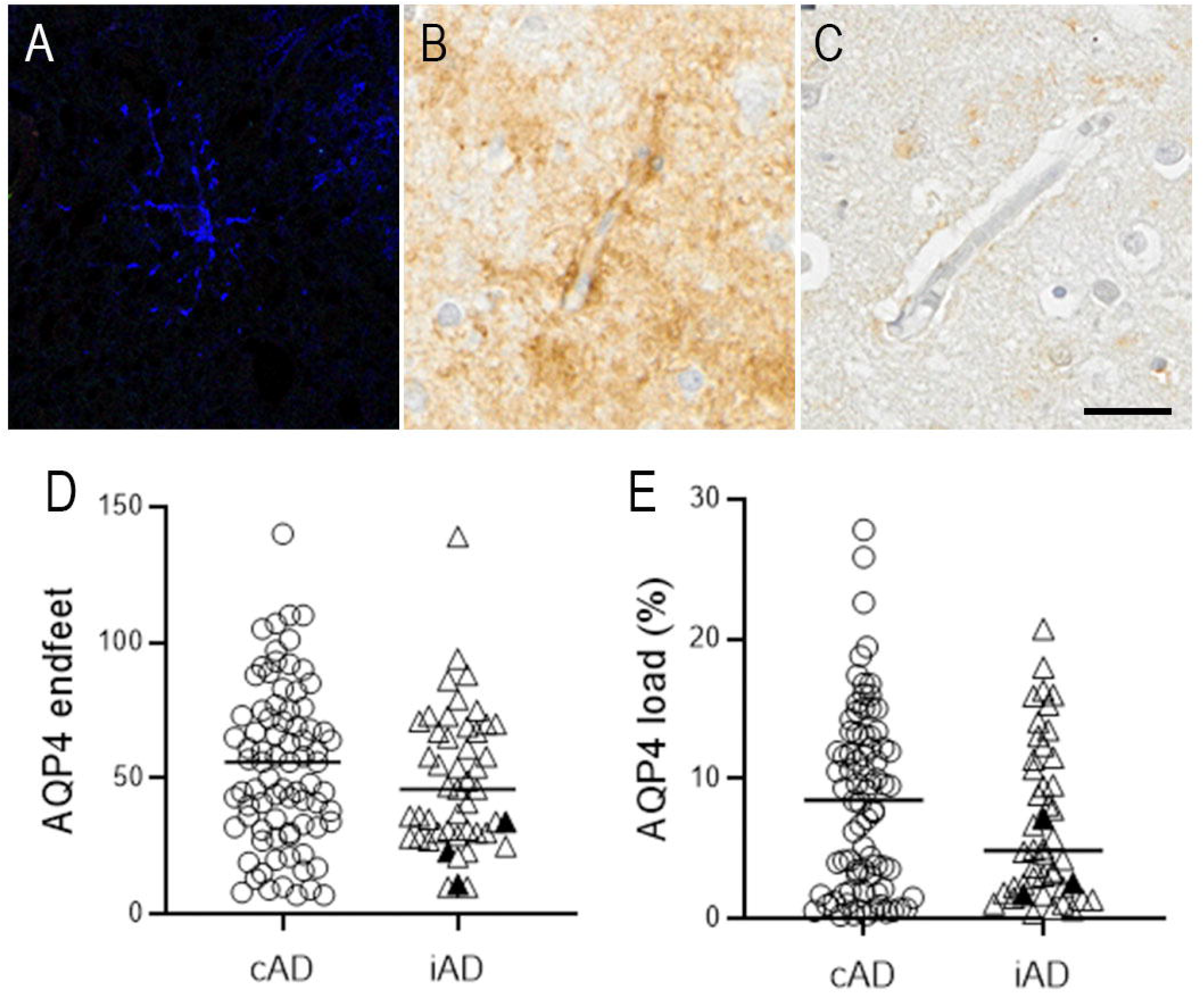
Aquaporin 4 (AQP4) expression in the cerebral cortex is present in both the cell body and processes of astrocytes (A). There is particularly prominent AQP4 staining around many (B) but not all (C) capillaries, representing AQP4 in astrocyte end feet. Quantification showed no significant difference between immunised AD cases (iAD) and control AD cases (cAD) in either the density of capillaries wrapped by AQP4-containing endfeet (D) or overall AQP4 load (E, % area stained). The case known to have had ARIA (case no. 1) is indicated in red. Scale bar = 50μm.

Quantification of peri-capillary AQP4-positive endfeet showed no significant difference between groups (median iAD=46 *vs* cAD=56, p=0.311, Figure 2D). Likewise, quantification of overall cortical AQP4 load showed no significant difference between the groups (median iAD=4.87% vs cAD=8.82%, p=0.294, Figure 2E). In the subgroup analysis in relation to *APOE* status, there was no difference in AQP4 load or AQP4 endfeet according to ε4 carrier status between the iAD and cAD groups (data not shown).

### The case with known ARIA-E (case no. 1)

Once the study was unblinded, detailed data were assessed for case no. 1, the only patient in the cohort who had imaging during life, in retrospect defined as having had ARIA-E [5, 7]. As previously reported, the case has extensive removal of Aβ and severe cortical and parenchymal CAA [5]. In this case (black triangle), quantification of capCAA, AQP4 endfeet and AQP4 load fell within the ranges for the other iAD cases (white triangles), not appearing strikingly different in this regard (figures 1H, 2D-E).

### Correlations

Correlations were assessed between capCAA, AQP4 endfeet data, AQP4 and Aβ42 loads and available clinical information.

In the iAD group, a significant negative correlation was observed between capCAA and AQP4 load (r = -0.498, p<0.001), and a trend towards an association between AQP4 load and capillary AQP4+ endfeet (r = 0.317, p=0.032) (Table 2A).

**Table 2:**
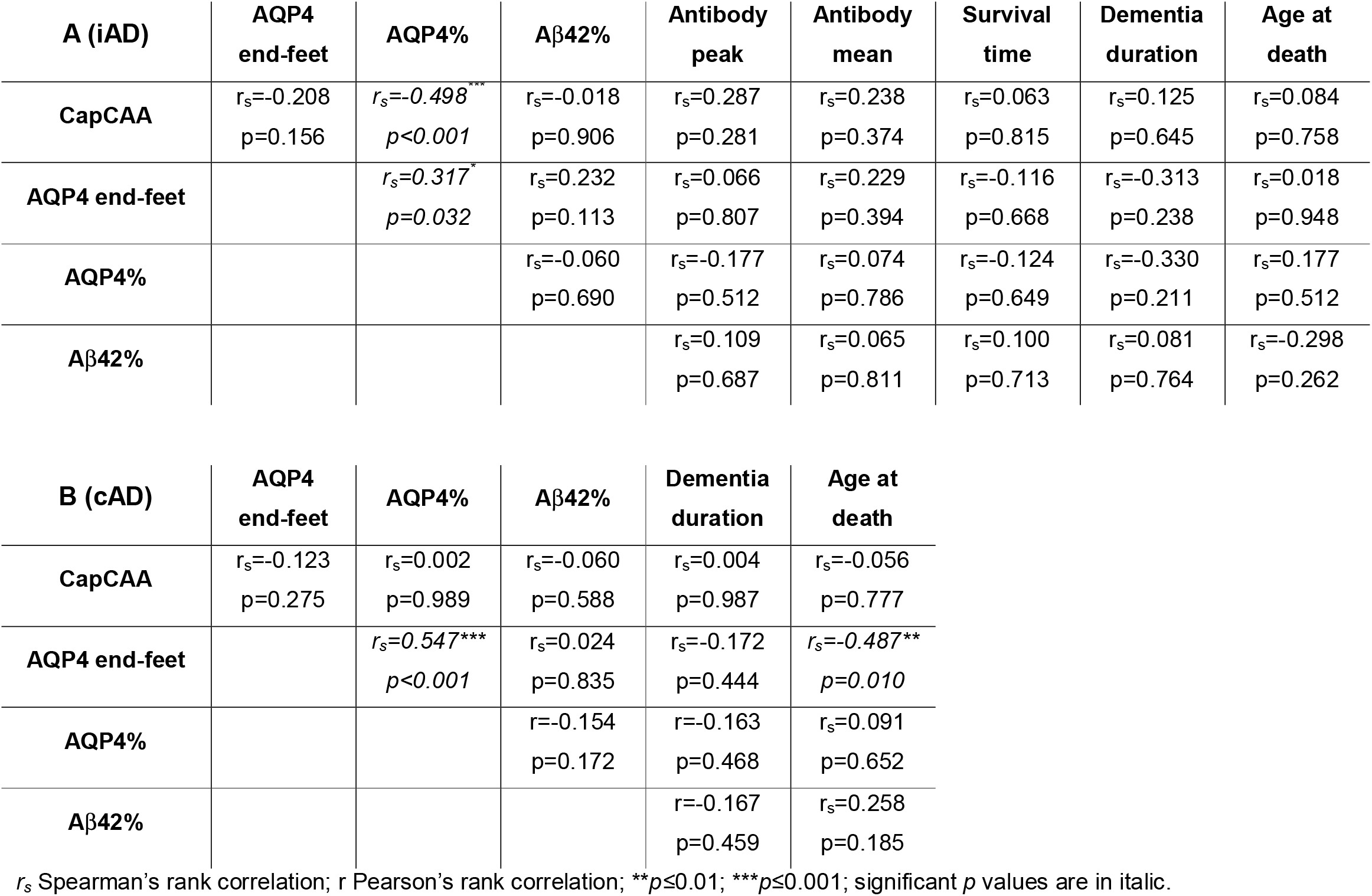
Correlations between the number of Aβ42+ capillaries (capCAA), AQP4+ capillaries (endfeet), overall AQP4 and Aβ42 load and the available clinical information in (A) the immunised AD group and (B) in the non-immunised AD group.

In the cAD cohort, a positive correlation was detected between the AQP4 load and AQP4+ endfeet (r = 0.547, p<0.001) and a negative correlation between AQP4+ endfeet and age at death (r = -0.487, p=0.010) (Table 2B). No other associations were observed in either group.

## Discussion

ARIA-E has been a persistent hurdle in clinical trials of Aβ immunotherapy for AD [19]. The presence of MRI signal changes, interpreted as reflecting oedema [16], have occurred in most clinical trials to date, causing significant dose reduction, interruption or trial cessation. Consequently, it is important to understand the pathophysiological basis for this problem, but understanding has been hampered by the paucity of neuropathological analyses of brain tissue from affected patients. There have been few autopsy studies of patients with AD in Aβ immunotherapy trials, other than the cases described here, and none with numbers of cases as large as the cohort investigated in this study. The impact of AN1792 immunotherapy on other aspects of AD pathology has been previously reported [5-9] - notably, the reduction in plaque load [5, 6, 47] and the increase in cerebral amyloid angiopathy (CAA) [5-8]. This has been attributed to Aβ from plaques being solubilised and drained towards the perivascular pathway (intramural periarterial drainage, IPAD) [13-15] as Aβ is cleared from the brain.

A similar increase in CAA severity has also been identified in animal models of AD immunised against Aβ including accumulation specifically in association with capillaries (capCAA). In AD mouse models, AQP4 the principal water transporter in the brain [32, 33, 48], is displaced away from capillaries following Aβ immunisation [31] – offering potential insight into the pathophysiology of ARIA-E. In this human study, we therefore investigated the impact of immunisation on capCAA and on the distribution of AQP4-positive astrocyte endfeet around capillaries.

### CapCAA

Prior to this study, the impact of immunisation on capillaries was unclear. As the capillary basement membrane is at the origin of the perivascular drainage pathway [12], it might be predicted that there would be increased capCAA following immunisation. In this study, we show that capCAA was 3.5 times higher in the immunised AD cohort, consistent with the hypothesis that Aβ from plaques is partly cleared via the perivascular drainage pathway and deposits in the capillary basement membrane forming capCAA [6, 8]. Interestingly, we did not see an association between capCAA and Aβ42 load in either of the groups. In AD, this might be explained by the very uncommon occurrence of capCAA (median of 1 event in 35 ROIs in AD) relative to the amount of Aβ in the form of plaques. Following immunotherapy, this might imply that only a small proportion of Aβ draining from plaques participates, resulting in the increased capCAA.

APOE4 is the largest genetic risk factor for sporadic AD [49, 50], with E4 homozygotes having a 90% lifetime risk of AD [51, 52]. The presence of ARIA-E has been strongly associated with possession of APOE4 – with each allele conveying 2.55x risk [16]. The risk of the APOE4 allele is thought to relate to the efficiency of APOE-mediated Aβ clearance [52], which may underpin the concurrent increase in capCAA. When we analysed the presence of capCAA according to APOE4 allele status in our cohorts, the E4 carriers in the immunised group had 8 times more capCAA than the non-E4 carriers; whereas no difference was observed in the unimmunised AD group when divided according APOE4 status. Consistent with our observation, two types of CAA have been defined, with type 1 including capCAA and associated with 4 times increased of the APOE4 allele frequency; while type 2 lacks capCAA and is mainly associated with APOE2 [25]. This implies that capCAA is more prone to form in E4 carriers, and CAA-Type 1 might be a contributory event underlying ARIA.

In addition, considering the fundamental role that capillaries play in maintaining cerebral homeostasis, for example in delivering oxygen and glucose and removing CO_2_, it is likely that disruption of the capillary structure by Aβ deposition will have deleterious consequences. CapCAA has been shown to cause capillary occlusion and functional blood flow disturbances [28], with other neuropathological features of AD such as tau deposits and neuroinflammation observed surrounding capCAA but not CAA [27]. Therefore, exacerbation of capCAA could potentially counteract a possible therapeutic benefit of lowered plaque burden after immunisation [4].

### AQP4

CapCAA may impact surrounding molecular and cellular structures, specifically perivascular astrocyte endfeet and associated AQP4 distribution. Indeed, displacement of AQP4 away from vascular Aβ with a concurrent increase of AQP4 around plaques has been reported previously in experimental models of AD [31, 40-42]. One study in human AD observed AQP4 staining associated with blood vessels in AD with mild CAA; while AD with moderate CAA had a diffuse pattern of AQP4, which was absent in AD with severe CAA [40]. In that study the staining was performed in n=3 per group and not quantified. In our larger and quantitative study, we did not observe changes in AQP4 load *per se* or in the number of capillaries surrounded by AQP4 after Aβ immunisation. However, a significant negative association was found between capCAA and AQP4 load, consistent with the idea that increased capCAA has led to alterations in AQP4 as demonstrated in mice [31].

### Implications for ARIA

A lymphocytic inflammatory reaction to the increased burden of Aβ in the vasculature, has also been proposed to play an important role in generating ARIA [15]. Close parallels have been noted between this post-immunotherapy iatrogenic change and the naturally occurring disorder of CAA-related inflammation [53, 54] in which patients produce anti-Aβ antibodies as an autoimmune response [55, 56]. Both CAA-related inflammation and severe CAA by itself are known to cause alterations on imaging of the underlying cerebral white matter, although the mechanism for this is unclear [57]. Both our own single case known to have ARIA and one other such published case [58] had a lymphocytic reaction in relation to the CAA and so inflammatory processes may have additional or greater relevance to ARIA than capCAA-associated alterations in AQP4.

A crucial limitation affecting all ARIA studies, ours notwithstanding, is the lack of neuropathological correlates. During the clinical phase of the AN1792 phase 1 trial, imaging was not routine and ARIA had not been described. In our cohort, one case retrospectively had imaging features consistent with ARIA-E (case no. 1) [5, 7], occurring nearly a year prior to death. As ARIA-E is a transient phenomenon, occurring 4 – 8 weeks after the first dose and lasting up to 113 days [20, 59], these changes may have fully resolved before death. This may explain why case no. 1 did not differ obviously from the other immunised AD cases in terms of capCAA, AQP4+ endfeet and AQP4 load. In addition, there may have been ARIA-E occurring in other cases in our cohort, that was not detected clinically or radiologically. Hence, it is challenging to be sure that we are looking at tissue affected by ARIA-E.

The best quality information about the relevant pathophysiological processes would be from biopsies or post-mortem samples from the time that ARIA occurred, but as far as we are aware no such samples exist. This is a substantial blind spot in the field of Alzheimer’s research. Understanding the mechanisms behind ARIA takes on a renewed level of importance given the recent approval by the Federal and Drug Administration (FDA) of Aduhelm (aducanumab), an anti-Aβ antibody, for the treatment of AD, with similar agents undoubtedly to follow. Aducanumab causes ARIA in clinical trials [60], and wider use may lead to more cases of ARIA. Especially given the controversy of whether aducanumab produces clinically meaningful improvement [61-63], we need a better understanding of ARIA to accurately appraise the balance of risks and benefits. Obtaining neuropathological correlates of ARIA should be a priority in the field.

### Conclusions

ARIA occurring in the context of Aβ immunotherapy in AD is hypothesised to result from dynamic changes in the localisation of Aβ with a shift of Aβ from plaques to blood vessel walls. This increased CAA severity is likely associated with inflammation and is analogous to the spontaneous disease of CAA-related inflammation. ARIA-E may be due to failure to control water flux at the blood-brain barrier. In support of this hypothesis, we found that Aβ immunotherapy is associated with increased in severity of capillary CAA, more so in carriers of APOE4 who are also at greater risk of ARIA. Therefore, exacerbation of capCAA could potentially counteract possible therapeutic benefit of lower plaque burden after immunisation, particularly in APOE4 carriers. Furthermore, capillary CAA is inversely related to AQP4 levels after immunisation, although we found no evidence of an overall change in AQP4 load or displacement of AQP4 from astrocyte endfeet. Limitations to our understanding of ARIA include the few cases from which tissue is available to study, lack of biopsies and difficulty in imaging of CAA and inflammation *in vivo*. The field must prioritise obtaining neuropathological correlates of ARIA to explore its mechanisms further.

## Data Availability

All data produced in the present study are available upon reasonable request to the authors.

## Acknowledgments

We would like to thank Dr Laura Palmer at the South West Brain Dementia Brain Bank (SWDBB) which is supported by BRACE (Bristol Research into Alzheimer’s and Care of the Elderly) and provided the AD cases. We thank Elan Pharmaceuticals for access to phase I AN1792 treatment study data and are grateful to all patients and carers who took part in the study. Our thanks also include the staff from Cardiff, Swindon, Oxford, Bath and Bristol Centres involved in the AN1792 study. We acknowledge the Histochemistry Research Unit and Biomedical Imaging Unit of the Faculty of Medicine, University of Southampton that facilitated tissue processing, staining and analysis.

## Ethical approval

The study was covered for the iAD cases by the ethical approval from Southampton and South West Hampshire Local Research Ethics Committees (Reference No: LRC 075/03/w) and for the AD cases from the South West Dementia Brain bank by the NRES Committee South West Central Bristol, REC reference: 08/H0106/28 + 5. All donors have given informed consent for autopsy and use of their brain tissue for research purposes.

## Authors’ contributions

CHH and KS immunolabelled, performed quantification and collected data. DAJ provided technical assistance. CHH and DB performed statistical analyses. CH advised on the clinical relevance of the findings. CHH and DB wrote the manuscript. DB and JARN conceived and designed the study. All authors read and approved the final manuscript.

## Availability of data and material

The dataset generated and/or analysed during the current study are available from the corresponding author on reasonable request.

